# Transcriptomic Similarities and Differences in Host Response between SARS-CoV-2 and Other Viral Infections

**DOI:** 10.1101/2020.06.18.20131326

**Authors:** Simone A. Thair, Yudong D. He, Yehudit Hasin-Brumshtein, Suraj Sakaram, Rushika Pandya, Jiaying Toh, David Rawling, Melissa Remmel, Sabrina Coyle, George N. Dalekos, Ioannis Koutsodimitropoulos, Glykeria Vlachogianni, Eleni Gkeka, Eleni Karakike, Georgia Damoraki, Nikolaos Antonakos, Purvesh Khatri, Evangelos J Giamarellos-Bourboulis, Timothy E Sweeney

## Abstract

COVID-19 is a pandemic that shares certain clinical characteristics with other acute viral infections. Here, we studied the whole-blood transcriptomic host response to SARS-CoV-2 and compared it with other viral infections to understand similarities and differences in host response. Using RNAseq we profiled peripheral blood from 24 healthy controls and 62 prospectively enrolled patients with community-acquired lower respiratory tract infection by SARS-Cov-2 within the first 24 hours of hospital admission. We also compiled and curated 23 independent studies that profiled 1,855 blood samples from patients with one of six viruses (influenza, RSV, HRV, ebola, Dengue, and SARS-CoV-1). We show gene expression changes in peripheral blood in patients with COVID-19 versus healthy controls are highly correlated with changes in response to other viral infections (r=0.74, p<0.001). However, two genes, *ACO1* and *ATL3*, show significantly opposite changes between conditions. Pathway analysis in patients with COVID-19 or other viral infections versus healthy controls identified similar pathways including neutrophil activation, innate immune response, immune response to viral infection, and cytokine production for over-expressed genes. Conversely, for under-expressed genes, pathways indicated repression of lymphocyte differentiation and T cell activation. When comparing transcriptome profiles of patients with COVID-19 directly with those with other viral infections, we found 114 and 302 genes were over- or under-expressed, respectively, during COVID-19. Pathways analysis did not identify any significant pathways in these genes, suggesting novel responses to further study. Statistical deconvolution using immunoStates found that M1 macrophages, plasmacytoid dendritic cells, CD14+ monocytes, CD4+ T cells, and total B cells showed change consistently in the same direction across all viral infections including COVID-19. Those that increased in COVID-19 but decreased in non-COVID-19 viral infections were CD56^bright^ NK cells, M2 macrophages, and total NK cells. The concordant and discordant responses mapped out here provide a window to explore the pathophysiology of COVID-19 versus other viral infections and show clear differences in signaling pathways and cellularity as part of the host response to SARS-CoV-2.

## Introduction

A novel coronavirus, SARS-CoV-2, has developed into a global pandemic, resulting in more than 7.6 million cases worldwide with over 427,000 deaths as we write (WHO accessed 14Jun2020)^1^. Contextually this pandemic is likely to surpass the SARS-CoV-1 2003 pandemic by 1000-fold whereby SARS-CoV-1 resulted in 8,098 cases, took 12 months to contain and had a 9.6% mortality rate (WHO accessed 1Jun2020). The novel SARS-CoV-2 virus, the causative agent for COVID-19 disease, is highly communicable and despite urgent and resource-intensive efforts globally, we have no vaccine or <u>efficacious</u> treatment in sight^2^.

COVID-19 clearly shares some immunological features with other viral responses, such as interferon activation, simultaneous repression of immune cells, and changes in metabolism including glucose and iron regulation as shown by cytokine and cytometry studies^3–5^. However, while many acute viral infections can lead to critical illness and death, COVID-19 appears both quantitatively and qualitatively to differ when compared to other acute viral infections. Notable features of COVID-19 include high rates of acute respiratory distress requiring mechanical ventilation; clinical coagulopathy; features of a cytokine storm and/or viral sepsis, and a high case fatality rate^6^. Thus, while studies comparing COVID-19 to healthy controls (HC) are useful, they do not explain the similarities and differences seen in the COVID-19 syndrome vs other viral infections.

Our approach involves a multi-cohort analysis of transcriptomic host response data to investigate host inflammation. The core discovery method leverages biological, clinical, and technical heterogeneity across datasets to identify generalizable disease biomarkers. We have repeatedly demonstrated that host response can be a generalizable sensitive and specific diagnostic and prognostic marker for presence, type, and severity of infections^7–9^, but also in autoimmune diseases, vaccination, TB, cancer and organ transplant^7,8,17–20,9–16^ We have shown in methodological work that this method produces results with the greatest reproducibility in independent cohorts^21^.

In this work, we used RNAseq to profile whole blood samples from 62 COVID-19 patients prospectively enrolled in Athens, Greece, together with 24 healthy controls. We simultaneously compiled a database of clinical viral infections from 23 studies of > 1,800 samples to represent the conserved immune response to a broad range of viral infections including influenza, RSV, HRV, SARS-CoV-1, ebola, and dengue. We here report on the results of a comparison of host responses to SARS-CoV-2 and other viruses. We mapped out their similarities and differences at the gene level, pathway level, and cell proportion level, as a first step to gain a better understanding of this novel pandemic virus.

## Methods

### SAMPLE ACQUISITION AND PROCESSING

#### COVID-19 samples from Hellenic Sepsis Study Cohort

A total of 76 adult patients with SARS-CoV-2 pneumonia were prospectively enrolled from April 1^st^ to May 4^th^ by department participating in the Hellenic Sepsis Study Group (www.sepsis.gr) using the inclusion criteria already described elsewhere^22^. Lower respiratory tract infection was defined as the presence of infiltrates in chest X-ray or chest computed tomography compatible with COVID-19. SARS-Cov-2 was detected by positive molecular testing of respiratory secretions. For patients who required mechanical ventilation (MV), blood sampling was performed within the first 24 h from MV. Exclusion criteria were infection by the human immunodeficiency virus, neutropenia, and any previous intake of immunosuppressive medication (corticosteroids, anti-cytokine biologicals, and biological response modifiers). The studies were conducted under the 30/20 approval by the National Ethics Committee of Greece. Written informed consent was provided by patients or by first-degree relatives in cases where patients were unable to consent.

Whole blood was drawn in PAXgene tubes at enrollment along with other standard laboratory parameters. Data collection included demographic information, clinical scores (SOFA, APACHE II), laboratory results, length of stay and clinical outcomes. Patients were followed up daily for 30 days; outcomes were defined as severe respiratory failure (PaO2/FiO2 ratio less than 150 requiring MV) or death. PAXgene Blood RNA samples were shipped to Inflammatix for processing.

#### Healthy control sample sourcing

Blood RNA tubes were prospectively collected from healthy controls (HC) through a commercial vendor (BioIVT) under IRB approval (Western IRB #2016165) using informed consent. Patients were non-febrile and verbally screened to confirm that no signs or symptoms of infection were present within 3 days prior to sample collection and that they were not currently undergoing antibiotic treatment, nor had not taken antibiotics within 3 days prior to sample collection. Furthermore, all samples were negative for HIV, West Nile, Hepatitis B, and Hepatitis C by molecular or antibody-based testing.

#### RNA extraction protocol

Prior to processing, samples in PAXgene Blood RNA tubes from 76 COVID-19 patients and 24 healthy controls were removed from -80C to thaw at room temperature for two hours. The samples were then inverted several times to achieve homogeneity, after which 3 mL aliquots were removed for processing. RNA was extracted from these samples using a modified version of the RNeasy Mini Kit (QIAgen) protocol executed on the a QIAcube automated workstation. PAXgene samples comprise of whole blood in PAXgene stabilizing solution. The sample is diluted with PBS, then centrifuged at 3,000 x g to pellet precipitated nucleic acids. Pellets were washed with molecular biology grade water and again pelleted via centrifugation at 3,000 x g. Pelleted material is resuspended in Buffer RLT (QIAgen). Using the automated QIAcube, samples are then subjected to treatment by Proteinase K and gDNA elimination via columns (QIAgen). Flow-through was mixed with isopropanol and passed over a MinElute (QIAgen) spin column. The column was washed with 80% ethanol and purified nucleic acid was eluted in RNase-free water. Purified RNA was heat denatured at 55° C for 5 minutes, then snap-cooled on ice. RNA was quantitated using a Qubit fluorimeter with the Quant-iT RNA Assay kit (Thermo-Fisher). Samples with an RNA integrity number (RIN) below 7 (BioAnalyzer, Agilent) did not proceed to sequencing, resulting in 62 COVID-19 samples and 24 HC samples for sequencing.

#### RNAseq library preparation

Total RNA samples were depleted of globin RNA using the GLOBINclear kit (Invitrogen) following the procedure described by the manufacturer. Globin-depleted RNA was quantified using the Qubit RNA High Sensitivity kit (Life Technologies) and 10ng of globin-depleted RNA was then used for rRNA depletion and RNAseq library preparation using the SMARTer Stranded Total RNAseq kit v2 Pico Input Mammalian (Takara Bio) following the manufacturer’s protocol. RNAseq libraries were then quantified using the Qubit dsDNA High Sensitivity kit (Life Technologies) and their quality and size evaluated by a Fragment Analyzer High Sensitivity Small Fragment kit (Agilent Technologies).

#### RNA sequencing

A total of 86 RNAseq libraries generated above were pooled and sequenced on an Illumina NovaSeq6000 Sequencing System (Illumina) in a paired-end fashion (2 × 100 cycles). 41 M to 124 M paired-end reads were obtained for each sample obtained for each sample. Fastq files were used as input for RNAseq data processing. Library prep and sequencing were performed at TB-SEQ (Palo Alto, CA).

### DATA PROCESSING AND ANALYSIS

#### RNAseq data processing

##### Trimming

Quality control (QC) assessment of the reads was done using FastQC ^23^. The adapter sequence and 3 bases on the 3’ end of the reads was trimmed using cutadapt as a commonly used procedure ^24^.

##### Alignment

Trimmed reads were mapped to a reference genome index generated based on the human genome, GRCh38, and a transcriptome reference, GENCODE v32 primary assembly gtf ^25^ with the sjdbOverhang option set to 100 (default), using STAR aligner (v2.7.3a).

##### Quantitation

Mapped reads were quantified as per Ensembl transcript ID as defined in GENCODE v32 annotation. Reads were summed across Ensembl transcript IDs mapping to Entrez gene IDs in order to compare them with other viral data assayed by microarrays (AnnotationDbi from Bioconductor)^26^.

##### Data Quality

Various QC metrics prior to and post trimming were examined to assess data quality as a standard procedure for RNAseq data. Additionally, the distributions of raw and trimmed counts were assessed and Principal Component Analysis (PCA) with various cutoffs was performed for QC. All 86 samples passed standard QC metrics and the resulting counts matrix (12,142 Entrez genes by 86 samples) was used in subsequent data integration steps (**Supplementary Table 1**).

#### Normalization and voom transformation of RNAseq counts

Low-expressed genes were filtered using the following cutoff: max counts per million (CPM) less than 5 across all 86 samples. Normalization factors were obtained using edgeR’s Trimmed Mean of M values (TMM) method^27^. The voom method was then used to transform counts into normalized log2-CPM **(Supplementary Figure 1)**^28^. Data is available at Gene Expression Omnibus (GEO) repository (GSE152641).

#### Non-COVID-19 viral dataset selection

Transcriptomic data of clinical respiratory infections caused by viruses other than SARS-CoV-2 were surveyed from Gene Expression OmniBus (GEO) and ArrayExpress for inclusion to define a conserved host response signature for non-COVID-19 viral infection. We identified 23 such independent datasets that profiled a total of 1,855 peripheral blood samples (PBMCs or whole blood) from patients (infants, children, or adults) with one of six viral infections (influenza, RSV, HRV, ebola, dengue, SARS-CoV-1, but not SARS-CoV-2). Collectively the 23 datasets comprised of 780 samples from healthy controls and 1,075 from patients with a viral infection represent biological, clinical, and technical heterogeneity observed in the real-world patient population with viral infections.

#### Non-COVID-19 viral dataset processing

Raw microarray data for each dataset was renormalized (when available) using standardized methods. Affymetrix arrays were renormalized using the robust multichip average (RMA) method. Illumina, Agilent, GE, and other commercial arrays were renormalized via normal-exponential background correction followed by quantile normalization. Data were log2-transformed. Probe to gene (Entrez ID) summarization was performed within each study using the mean signal intensity for probes mapping to a single gene.

#### COCONUT conormalization of all data sets

Of the 23 non-COVID-19 viral infections datasets, 20 datasets with a total of 879 viral infected patients and 754 HCs met the criteria for conormalization: 1) the dataset must have HCs, and 2) the dataset was obtained on a single-channel microarray platform. Integrated with the voom-transformed RNAseq dataset for COVID-19, they were conormalized together using COCONUT as previously described^8^. COCONUT uses COMBAT empiric-Bayes conormalization on healthy controls to derive correction factors for diseased patients. The technique integrates datasets such that (i) no bias is introduced to the diseased samples, (ii) there is no change to the distribution of a gene within a study, and (iii) each gene shares the same distribution across healthy controls between studies after normalization. This COCONUT conormalized expression data comprising of a total of 941 (COVID-19 and non-COVID-19) viral patients and 778 HCs across 9,818 genes common across 11 platforms were used as input data to perform the following multicohort and integrated analyses.

#### COVID-19 versus healthy control comparison

Hedges’ g effect size (ES)^29^ for each gene was calculated for COVID-19 (62) versus HC (24) two-group comparison test from the COCONUT conormalized output. P-value was calculated using a student’s t-test and adjusted using the Benjamini-Hochberg method to obtain the False Discovery Rate (FDR). ES threshold of ≥ 1 or ≤ -1 in combination with FDR threshold of ≤ 0.05% was used to identify genes whose expressions are over- or under-expressed in COVID-19 infected patients than in the mean value of HCs.

#### Non-COVID-19 viral versus healthy controls comparison

14 datasets composed of 1,324 whole blood and PBMC samples were chosen for the discovery cohort, of which 652 were from respiratory viral infected patients (viral) and 672 samples were from HCs patients. As a multi-cohort analysis with conormalized data as input, we utilized a well-established MetaIntegrator (version 2.1.1) as described previously^30^. Briefly, Hedges’ g ES was computed for each gene within a study between viral and HC. ESs for genes across studies was summarized using the DerSimonian& Laird random-effects model, where each ES is weighted by the inverse of the variance in that study^31^. We used an ES threshold ≥ 1 or ≤ -1 with FDR ≤ 0.05% to identify signature genes (**Supplementary Table 2**).

#### Validation of non-COVID-19 viral infection signature

The signature genes identified based on 14 discovery datasets were evaluated for prediction of viral infections from HC with a score calculated for each sample using the following formula:

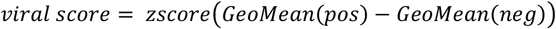

The score is a rescaled difference between geometric means of positive (over-expressed) genes and negative (under-expressed) genes. Receiver-operating characteristics (ROC) plots are generated for held out validation datasets and the Area Under the ROC (AUC) is used as a performance metric. For validation of the non-COVID-19 viral signature, we compiled 9 datasets comprised of 6 held out from the COCONUT expression data, plus 3 normalized as per platform requirements without COCONUT (**Table 3**). We then tested this signature first using 4 datasets comprising of 178 respiratory viral infection samples and 58 HCs (236 total) (**Table 3**). We then further validated this signature in 5 datasets of other viral etiology (245 viral and 50 HC, 295 total) **(Table 3)**.

#### COVID-19 versus non-COVID-19 viral Comparison

Hedges’ g ES was calculated for each gene in a COVID-19 (62) and non-COVID-19 viral (652) two-group comparison test from the COCONUT conormalized expression data. P-value was calculated using a Welch’s t-test assuming unequal variance and sample sizes and adjusted using the Benjamini-Hochberg^32^ method to obtain the False Discovery Rate (FDR). ES threshold ≥ 1 or ≤ -1 in combination with FDR threshold of ≤ 0.05% was used to identify signature genes.

### PATHWAY AND IMMUNOSTATES ANALYSIS

#### Pathway Analysis

Each over- or under-expressed gene set from comparisons between COVID-19 vs HC, non-COVID-19 viral infection vs HC, and COVID-19 vs non-COVID-19 viral infection was subjected to a pathway analysis with Gene Set Enrichment Analysis^33^. We tested significance of over-representation of genes in each of the pathways reflected in Gene Ontology (GO) including biological process (BP), molecular function (MF), and cellular compartment (CC). The human transcriptome reference is used as background and the p-values from the hyper-geometric test were adjusted using the Benjamini-Hochberg method ^32^. Top-ranked pathways common between COVID-19 and non-COVID-19, and specific separately to COVID-19 or non-COVID-19 viral infections were selected.

#### ImmunoStates Analysis

A statistical deconvolution method was used to estimate the percentage of 25 immune cell types in the peripheral blood transcriptome data ^34,35^. Statistical deconvolution estimates the percentage of various cell types present in a blood transcriptome profile. It uses a set of pre-defined genes that represent cell types of interest, called a basis matrix, and a variant of linear regression to make estimates. Previously, it was demonstrated that different methods produce highly correlated estimates of cellular proportions once basis matrix is fixed^34^. Here, immunoStates (MetaIntegrator) was used as a basis matrix because it has been shown to reduce the effect of the biological and technical heterogeneity in transcriptome data on statistical deconvolution and identify robust changes in immune cell proportions ^34–37^. The 14 non-COVID-19 viral discovery datasets and the COVID-19 dataset were deconvolved separately, then change in proportion of a given cell type between healthy controls and the infected patients of each dataset was estimated.

## Results

### Differential expression analysis of transcriptome profiles of patients with COVID-19

We prospectively enrolled and sequenced RNAseq from whole blood from 62 patients with COVID-19 and 24 healthy controls (**Table 1)**. Differential expression analysis of 86 peripheral blood samples identified 2,002 differentially expressed genes (771 over-expressed, 1,231 under-expressed; **Figure 1a, Supplementary Table 2**) with absolute ES ≥ 1 and FDR ≤ 0.05%), referred to as <u>COVID-19 signature.</u> We performed pathway enrichment analysis of the COVID-19 signature using Gene Ontology (GO) terms. The 30 most significant pathways for 771 over-expressed genes included neutrophil activation, innate immune response, immune response to viral infection, type-I interferon signing and cytokine production (**Figure 1b**), and for 1,231 under-expressed genes include lymphocyte differentiation and T cell activation and regulation (**Figure 1c**). These results suggest that in response to SARS-CoV-2 infection T cells are suppressed whereas neutrophils are activated as a hallmark of its overwhelming host response represented in the transcriptomic changes. High neutrophil-to-lymphocyte ratios have been observed as a marker of severity in sepsis, cancer, and pneumonia^38–41^.

**Figure 1.**
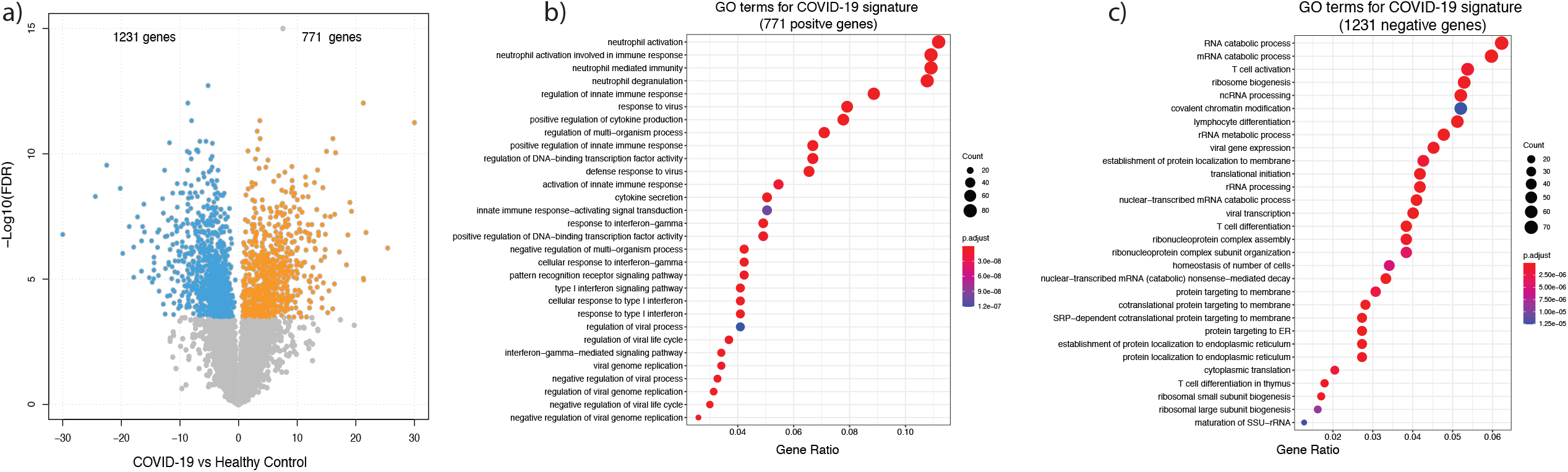
RNA-seq data of COVID-19 patients vs healthy control. (a) Significance score [defined as -log10(FDR)] vs mean difference of co-normalized log2-transformed expression data between COVID-19 patients (n = 62) vs healthy controls (n = 24). The chosen cutoff of ES ≥ 1 or ≤ -1 with FDR ≤ 0.05% yields the <u>2,002 COVID-19 signature</u>, including 771 positively regulated genes and 1,231 negatively regulated genes. GO term enrichment analysis of positive (b) and negative (c) gene sets reveal increased neutrophil function enrichment and decreased T cell related pathways (Gene Ratios represent the number of genes in our gene set within that pathway).

**Table 1.** Baseline characteristics table for COVID-19 patients. All continuous variables are reported as median and interquartile ranges [IQR] (n).

|  | COVID-19 Patients |
| --- | --- |
| n | 62 |
| Age in years: median [IQR] (n) | 61 [52,70] (61) |
| Gender = Male (%) | 40 (65) |
| SOFA score | 2 [1,4] (61) |
| APACHE II | 6.5 [4,9] (56) |
| Pneumonia severity index | 89.5 [65,104.5] (48) |
| White blood cell (mm3) | 6180 [4910,8420] (59) |
| Neutrophils | 75.5 [65.43,84.13] (59) |
| Lymphocytes | 15.69 [10.5,22.55] (59) |
| Platelets (k/ mm3) | 195.2 [158.8, 238.8] (58) |
| Lactate (mmol/l) | 1.55 [1.04,2.08] (30) |
| pO2.FiO2 (mmHg) | 255.35 [112.5,310.8] (50) |
| Creatinine (mg/dl) | 0.9 [0.7,1.015] (58) |
| PCT (ng/ml) | 0.1 [0.04,0.41] (49) |
| CRP (mg/l) | 78.85 [29.48,175.8] (60) |
| Days btwn onset symptoms and sampling | 6 [4,8] (53) |
| Days btwn intubation and sampling | 1 [0.5, 1.5] (23) |
| Days btwn hospital admission and intubation | 2 [1, 3.5] (23) |

### Identification of host response genes to viral infections through multi-cohort analysis

Based on our previous results ^14^, we hypothesized that there is a conserved immune response to respiratory viral infections irrespective of age and genetic background of a patient or a virus. We identified 23 studies of acute viral infection, and from these selected 14 as our discovery set for a non-COVID-19 viral signature (**Table 2**), and 9 were held out for validation. Statistical power analysis^42^ found that even with high inter-study heterogeneity, we had more than 80% statistical power at p-value = 0.01 for detecting absolute ES > 0.43 in these datasets (**Supplementary Figure 2**). The multi-cohort analysis of 1,324 transcriptome profiles (652 non-COVID-19 viral patients, 672 healthy controls) from these 14 studies using MetaIntegrator^30^ identified 635 differentially expressed genes (314 over-expressed, 321 under-expressed). ROC plots for all of the discovery datasets using this signature illustrate the high sensitivity and specificity this gene list possesses, indicating genes that are highly discriminatory and hence likely to represent this conserved signature **(Figure 2a, Supplementary Table 2**). We refer to these 635 genes in short as the <u>non-COVID-19 viral signature.</u> Similar to the COVID-19 signature, GO analysis of over- and under-expressed genes in the non-COVID-19 viral signature identified a similar set of pathways highlighted by neutrophil and T cell activation, respectively (**Figure 2b, 2c**).

**Figure 2.**
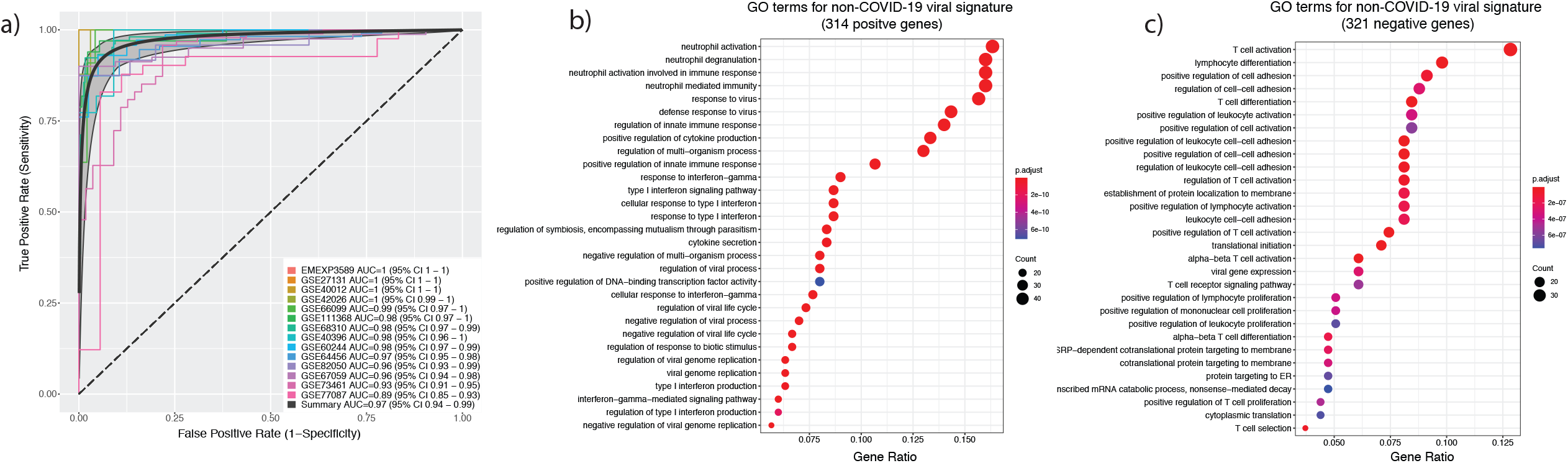
MetaIntegration of 14 non-COVID-19 viral disease datasets. (a) Multicohort analysis of 14 datasets of viral infections (n = 652) and healthy controls (n = 672) identified <u>635 non-COVID-19 viral signature</u>, including 314 positively regulated genes and 321 negatively regulated genes at the chosen cutoff of ES ≥ 1 or ≤ -1 with FDR ≤ 0.05%. GO term enrichment analysis of positive (b) and negative (c) gene sets reveal increased neutrophil function enrichment and decreased T cell related pathways, similar to those in Figure 1 (Gene Ratios represent the number of genes in our gene set within that pathway).

**Table 2.**
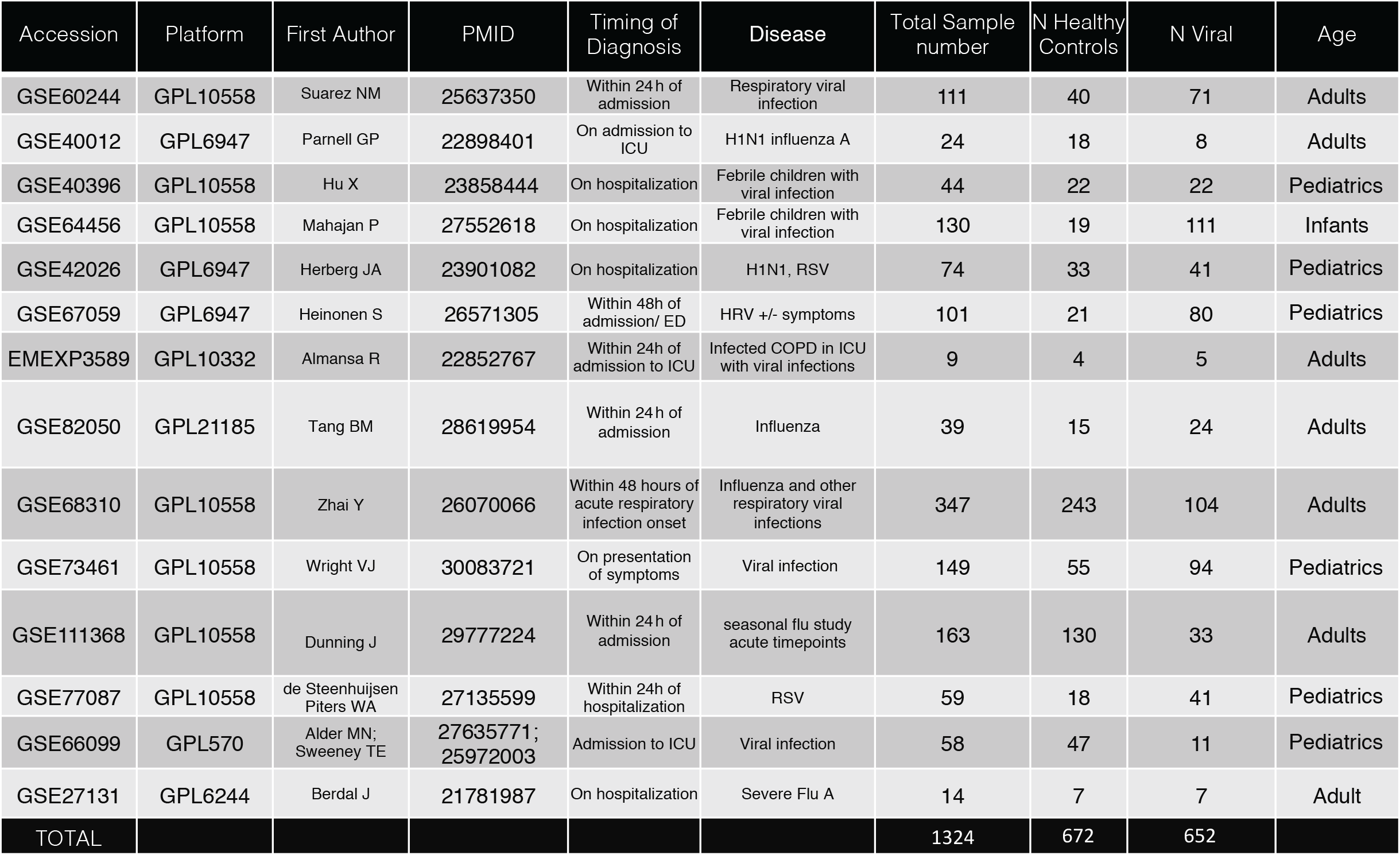
14 datasets used for discovery of the non-COVID-19 viral immune response.

### Validation of host response genes to viral infections in multiple independent datasets

Next, we confirmed that the non-COVID-19 viral signature is conserved across viruses by validating it in several independent datasets. We calculate the non-COVID-19 viral score for a sample as the difference in geometric means of over-expressed and under-expressed genes. In four independent studies consisting of 236 samples (178 viral infections, 58 healthy controls; **Table 3**), the score accurately distinguished patients with a respiratory viral infection (influenza, HRV, or RSV) from HCs (**Figure 3a**).

**Figure 3.**
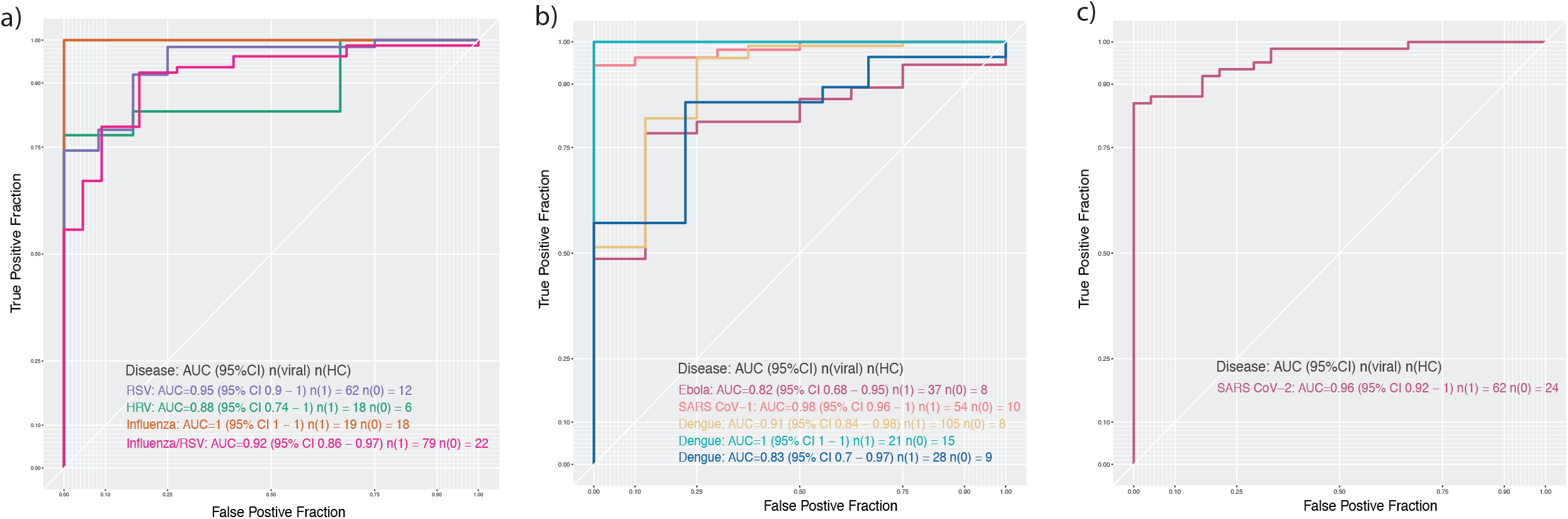
Validation of a global host immune response to viral infections. (a) ROC performance of <u>635 non-COVID-19</u> signature in 4 independent datasets including HRV, RSV, picornavirus and influenza. (b) ROC performance in 5 additional cohorts of disease not included in discovery [Ebola (GSE122692), SARS CoV-1 (GSE5972) and Dengue (GSE38246, EMTAB3162, GSE51808)]. (c) The signature is also tested in the 62 COVID-19 patients and 24 HCs.

**Table 3.** Datasets for validation of the non-COVID-19 viral vs healthy signature. * indicates datasets not eligible for COCONUT

| Accession | Platform | First Author | PMID | Timing of Diagnosis | Disease | Total Sample number | N Healthy Controls | N Viral | Age |
| --- | --- | --- | --- | --- | --- | --- | --- | --- | --- |
| GSE117827 | GPL23126 | Yu J | 30339221 | Within 24h of hospitalization | HRV | 24 | 6 | 18 | Pediatric |
| GSE20346 | GPL6947 | Parnell G | 21408152 | At peak symptoms | Influenza | 37 | 18 | 19 | Unknown |
| GSE34205 | GPL570 | Ioannidis I | 22398282 | Within 42-72h of hospitalization | Influenza/RSV | 101 | 22 | 79 | Pediatric |
| GSE103842 | GPL10558 | Rodriguez-Fernandez R | 29045741 | Within 24h of hospitalization | RSV | 74 | 12 | 62 | Pediatric |
| TOTAL |  |  |  |  |  | 236 | 58 | 178 |  |

**Table 3.** Datasets for validation of the non-COVID-19 viral vs healthy signature.
| Accession | Platform | First Author | PMID | Timing of Diagnosis | Disease | Total Sample number | N Healthy Controls | N Viral | Age |
| --- | --- | --- | --- | --- | --- | --- | --- | --- | --- |
| GSE5972 | GPL4387 | Cameron MJ | 17537853 | Within 24h of hospitalization | SARS (CoV1) * | 64 | 10 | 54 | Adults |
| GSE122692 | GPL16686 | Reynard S | 30626757 | Within 24h of hospitalization | Ebola * | 45 | 8 | 37 | Adults |
| EMTAB3162 | GPL570 | van de Weg CA | 25768297 | On admission | Dengue | 36 | 15 | 21 | Adults and Pediatric |
| GSE51808 | GPL13158 | Kwissa M | 24981333 | On admission | Dengue | 37 | 9 | 28 | Adults and Pediatric |
| GSE38246 | GPL15615 | Popper SJ | 23285306 | Within 24h of hospitalization | Dengue * | 113 | 8 | 105 | Pediatric |
| TOTAL |  |  |  |  |  | 295 | 50 | 245 |  |

Second, we investigated whether the non-COVID-19 viral signature is observed in other severe viral infections including ebola, dengue, and SARS-CoV-1 in five independent studies (50 HC, 54 SARS-CoV-1, 37 ebola, 154 dengue). In each study, the non-COVID-19 viral score also distinguished patients with a viral infection from healthy controls with high accuracy (**Figure 3b**).

Third, we tested whether the non-COVID-19 viral signature would also distinguish patients with COVID-19 from healthy controls. We calculated the non-COVID-19 viral score for each of 62 COVID-19 patients together with 24 HCs using the conormalized expression data. We found that non-COVID-19 viral score separated patients with COVID-19 from HCs with an AUC of 0.96 (**Figure 3c**), similar to SARS-CoV-1 (AUC=0.98).

### Comparison of COVID-19 profile with non-COVID-19 viral infection profile

Next, we investigated similarities and differences in host response to SARS-CoV-2 and other respiratory viruses by comparing change in expression with respect to healthy controls across 9,818 genes that were present across all datasets. When considering the entire transcriptome, there was high correlation (r = 0.74, p < 0.001) between change in expression in response to SARS-CoV-2 or other respiratory viruses (ES from COVID-19 vs HC comparison is plotted against ES from non-COVID-19 vs HC comparison in **Figure 4a**). We visualized *<u>2,002 COVID-19 signature genes</u>* and *<u>635 non-COVID-19 signature genes</u>* in the same ES scatter plot by different colors to highlight their relationships (**Figure 4a, Supplementary Table 2**). We observe that 7,626 genes uncolored in the middle (gray, with higher density in the center shown by contours) out of 9,818 profiled (77.7%) are not in the signature genes in either COVID-19 or non-COVID-19 viral infections. Given the high correlation (r = 0.74), it is not surprising that 223 genes are concordantly over-expressed (ES ≥ 1, FDR ≤ 0.05%) as well as 220 genes concordantly under-expressed with (ES ≤ -1, FDR ≤ 0.05%). Of the remaining genes from the *<u>non-COVID-19 signature</u>*, there are 90 genes over-expressed and 100 genes under-expressed in non-COVID-19, however these had ES between -1 and 1 in the distribution of the COVID-19 ESs. As well, of the remaining genes from the *<u>COVID-19 signature</u>*, there are 547 genes over-expressed and 1,010 genes under-expressed in COVID-19 that had ES between -1 and 1 in the distribution of the non-COVID-19 ESs. We only found two genes that were completely discordant, thus completely oppositely regulated in COVID-19 and non-COVID-19 viral infections: Aconitase1 (*ACO1*) over-expressed in COVID-19 and under-expressed in non-COVID-19 viral infections and Atlastin GTPase 3 (*ATL3*) over-expressed in non-COVID-19 viral infections and under-expressed in COVID-19. Interestingly, *ACO1* is involved in iron metabolism, and heme appears to be interlinked with COVID-19 pathophysiology^43^. *ATL3* is required for endoplasmic reticulum (ER) membrane junctions and may be linked to viral replication sites^44^.

**Figure 4.**
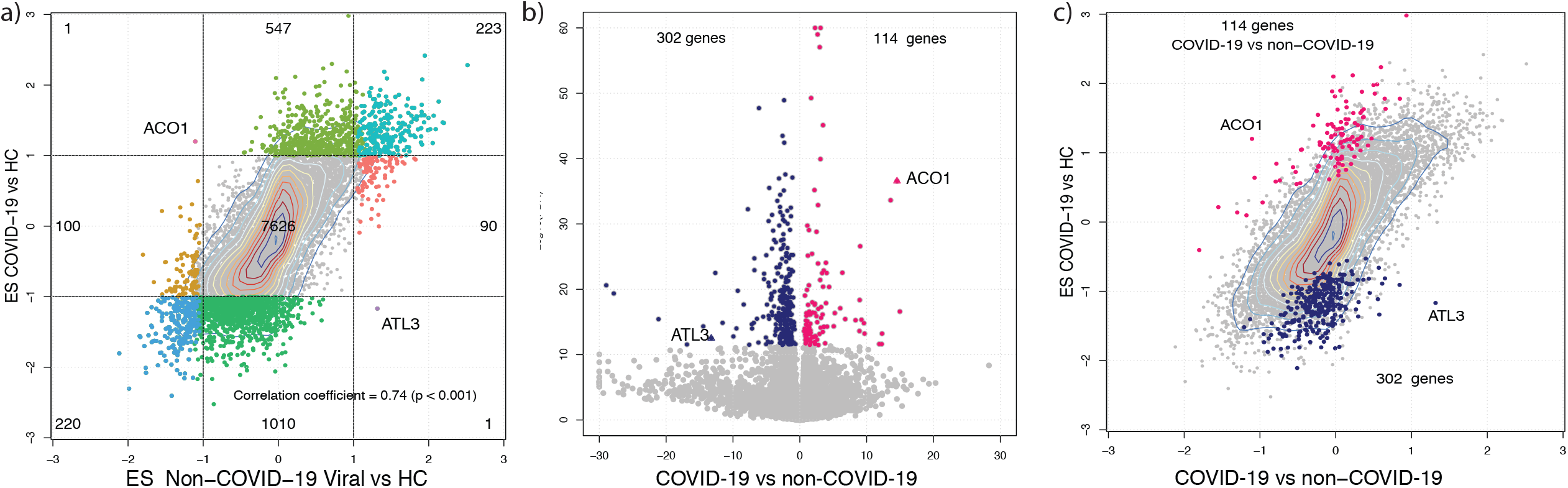
Comparison of COVID-19 signature with non-COVID-19 signature. (a) Scatter plot of effect size for all 9,818 genes commonly present in all datasets between non-COVID-19 vs HC (x-axis) and COVID-19 vs HC (y-axis). <u>2,002 COVID-19 signature</u> genes from Figure 1 and <u>635 non-COVID-19 signature</u> genes from Figure 2 are colored distinguishably in 9 quadrats. Concordant host response between COVID-19 and other viral infections is reflected by 223 commonly positively and 220 negatively regulated genes in both. Discordant response is only seen in ACO1 whose expression is positively regulated in COVID-19 but negatively regulated in non-COVID, and in ATL3 whose expression is negatively regulated in COVID-19 but positively regulated in non-COVID-19. (b) The head-to-head comparison between COVID-19 and other viral infections was made possible by using co-normalized data by COCONUT. Significance score [defined as -log10(FDR)] vs mean difference of co-normalized log2-transformed expression data between COVID-19 patients (n = 62) vs other viral infections (n = 652). The chosen cutoff of ES ≥ 1 or ≤ -1 with FDR ≤ 0.05% yields <u>416 COVID-19 specific signature,</u> including 114 positively regulated genes and 302 negatively regulated genes. (c) To illustrate the overlap of (a) and (b), the <u>416 COVID-19 specific signature</u> genes from head-to-head comparison in (b) are shown in the same scatter plot in (a).

Therefore, in order to identify a statistically significant set of genes differentially expressed in COVID-19 patients compared to those with other viral infections, we employed COCONUT to conormalize the two disease types into a single matrix for comparison of 62 COVID-19 patients versus 652 non-COVID-19 viral infection patients. Using COCONUT allows for comparison across datasets with heterogeneity while simultaneously creating a way to calculate an FDR for the gene effect size when compared “head-to head” or “disease to disease” directly, rather than looking for correlated and anti-correlated genes for which ES and FDR are calculated separately. At |ES| ≥ 1 with FDR ≤ 0.05%, we found 416 genes as <u>COVID-19-specific genes</u>, 114 over-expressed and 302 under-expressed in patients with COVID-19 than in those with non-COVID-19 viral infection **(Figure 4b**). To illustrate the gain in identification of genes to investigate and re-iterate the value in this statistical method, this set of genes from (b) are highlighted in the same scatter plot from panel a **(Figure 4c)**.

Unlike the COVID-19 and non-COVID-19 viral signatures, the pathway analysis of this gene set did not identify any statistically significant GO terms, potentially indicating novel pathophysiology unique to COVID-19. This combination of genes may include those less well annotated within pathways and thus less likely to result in statistically significance assignment to a pathway. Nonetheless, top-ranked but statistically insignificant GO terms include muscle contraction, regulation of epithelial cell proliferation, and biological processes involved in lung and respiratory development for 114 positive genes, as well as pathways related to T cell homeostasis and T cell differentiation for 302 negative genes. The significance of these pathways in connection with clinical manifestation needs to be investigated further.

### Similarities and differences in pathways between COVID-19 and non-COVID-19 viral infection

We expanded our comparison of significant pathways in response to SARS-CoV-2 versus non-COVID-19 viruses by including all pathways instead of only 30 most significant pathways. We found pathways for over-expressed genes are highly concordant between patients with COVID-19 and non-COVID-19 viral infections (**Figure 5a**), pathways for under-expressed genes are discordant (**Figure 5b**).

**Figure 5.**
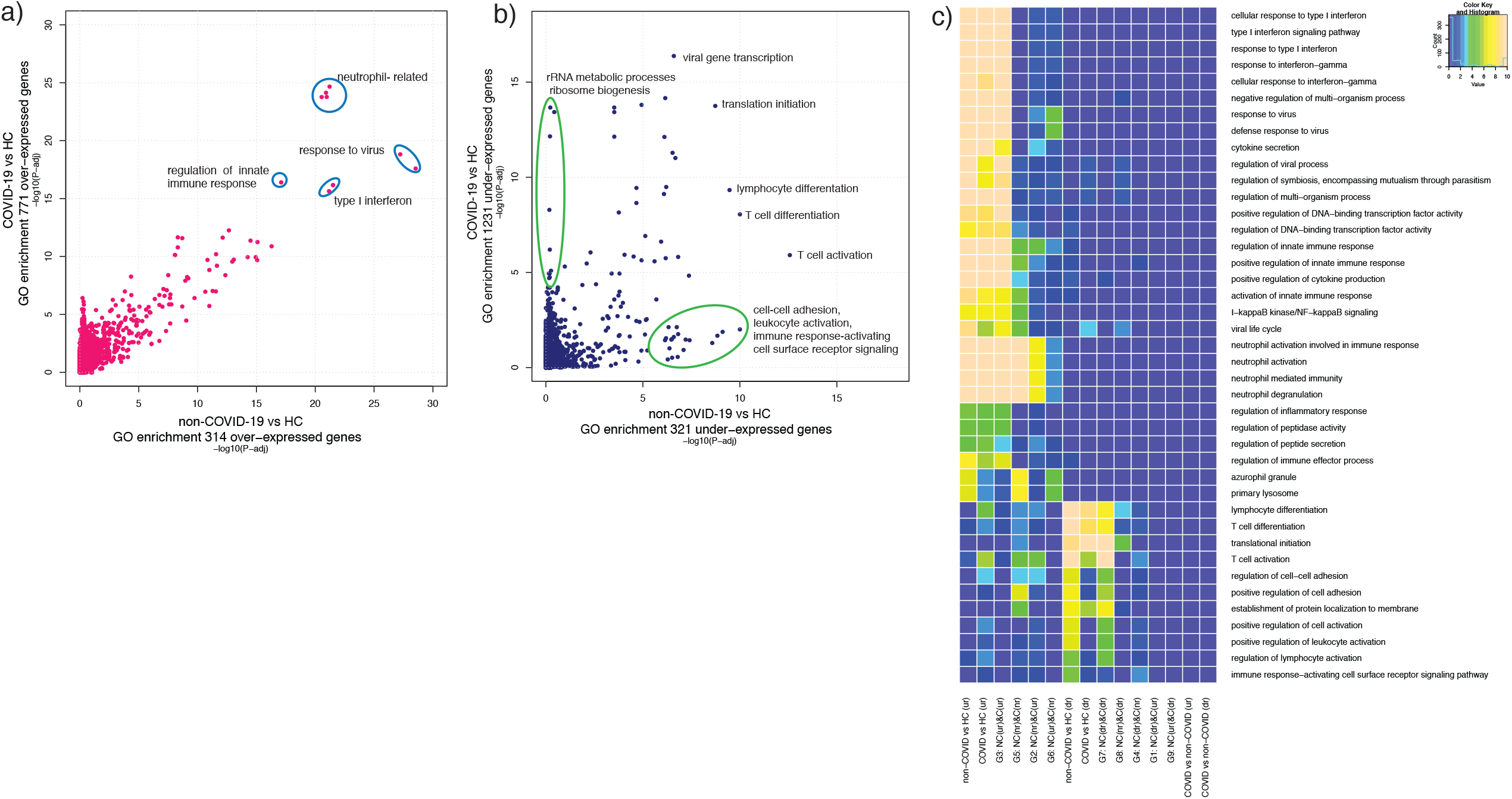
Summary of pathway analysis results. Scatter plots of the significance level from pathway enrichment analysis between COVID-19 and non-COVID-19 viral infections obtained for positive genes in (a) and negative genes in (b) respectively. The significance level is defined as -log10(BH-corrected p-value) for each pathway. The concordance is seen in results for up-regulated genes between COVID-19 and non-COVID-19, while a degree of discordance is evident in down-regulated genes between COVID-19 and non-COVID-19. (c) The heatmap shown as the significance level in each gene set of interest including COVID-19 vs HC (+) and (-), non-COVID-19 viral vs HC (+) and (-), and COVID-19 vs non-COVID-19 viral (+) and (-), together with 8 gene sets from Figure 4a (column) belonging to a pathway represented by Gene Ontology (row). Scale in heatmap is from 1 to 10 for the significance level.

To amalgamate these findings we performed hierarchical clustering of all pathway analysis results of all gene sets of interest including *three signature sets*: 1) COVID-19 vs HC (771 over-and 1,231 under-expressed), 2) non-COVID-19 viral vs HC (314 over- and 321 under-expressed), and 3) COVID-19 vs non-COVID-19 viral (114 over- and 302 under-expressed) as well as the *8 gene groups* based on concordance between signatures **(Figure 5c, Supplementary Table 2**). To check the dependency of GO term enrichment results on the cutoffs for selecting signature genes, we tested three additional cutoffs (less or more stringent than the chosen one) each for COVID-19 vs HC, non-COVID-19 vs HC, or COVID-19 vs non-COVID-19 comparison. The results for over-expressed, under-expressed, and all genes from each cutoff together with the 9 gene sets from **Figure 4a** show a merging and comprehensive picture of pathway analysis results **(Supplementary Table 3, Supplementary Figure 4**) allowing one to focus on pathways of interest, either commonly significant across gene sets or uniquely significant in a gene set or a combination of genes of interest.

### Similarities and differences in changes in immune cell proportions between COVID-19 and non-COVID-19 viral infection

We estimated proportions of 25 immune cell types in bulk gene expression in blood samples from patients with COVID-19 or non-COVID-19 viral infections using immunoStates. In patients with COVID-19, we found immune cells from myeloid-lineage (M1 macrophages, neutrophils, and MAST cells) increased significantly (FDR ≤ 10%), and lymphoid cells (CD4+ and CD8+ alpha-beta T cells, B cells) decreased significantly (FDR ≤ 10%) during viral infection (**Figure 6a, Supplementary Table 3**). These results are in line with recent reports demonstrating increased neutrophil and decreased T cell counts in COVID-19 patients^39–41^. In patients with non-COVID-19 viral infections, we observed significant increase in proportion for myeloid cells (M1 macrophages, CD14+ monocytes, MAST cells), and significant decrease in proportion for lymphoid cells (CD4+ and CD8+ T cells, gamma-delta T cells, B cells) (**Figure 6b, Supplementary Figure 3)**. Indeed, when considering changes within each dataset, M1 macrophages, plasmacytoid dendritic cells, CD14+ monocytes, CD4+ T cells, and total T cells showed change consistently in the same direction across all viral infections including COVID-19 (**Figure 6b**).

**Figure 6.**
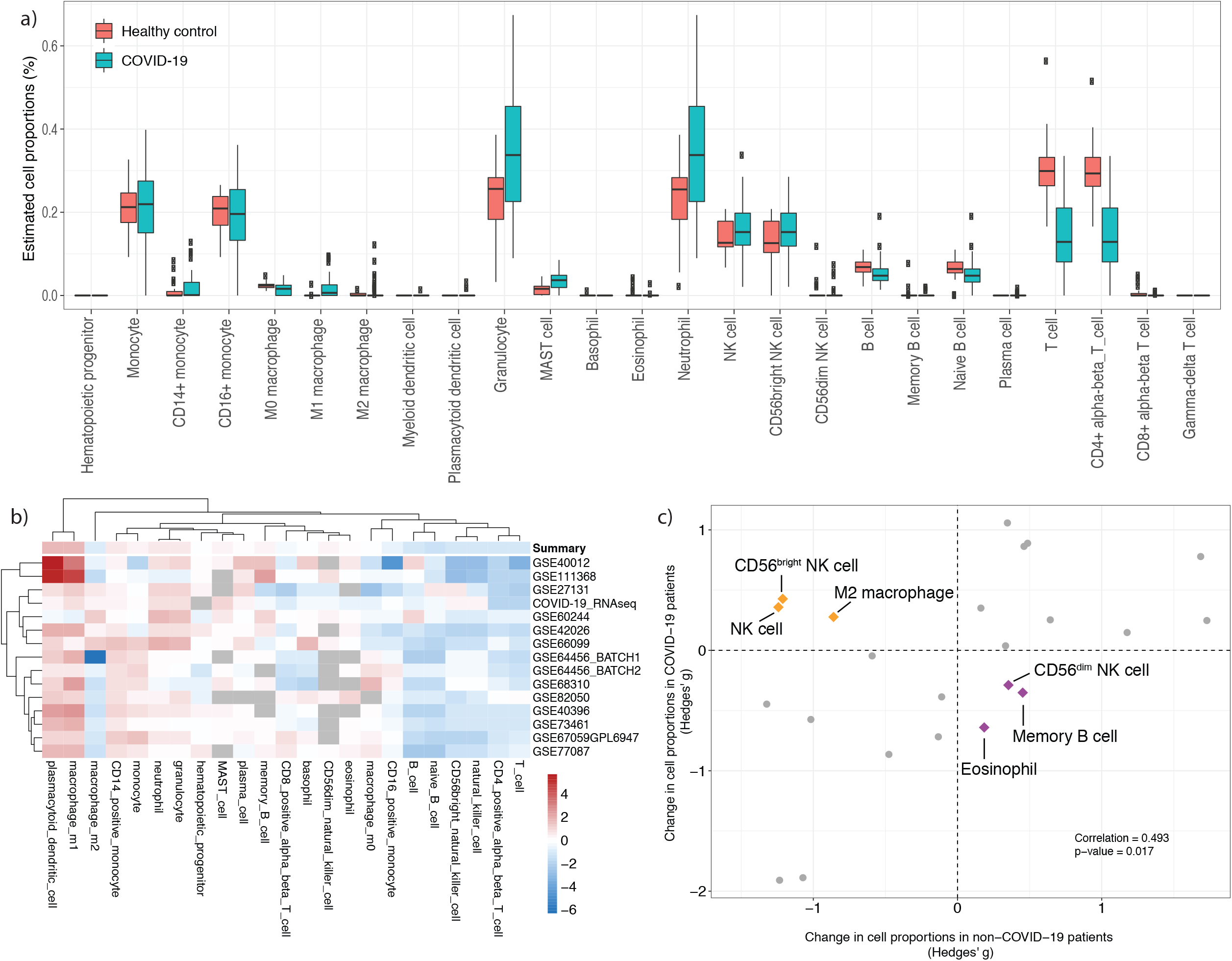
Statistical deconvolution of bulk transcriptome profiles using immunoStates of COVID-19 versus non-COVID-19 viral infections. a) Changes in cell proportions when comparing COVID-19 patients to healthy controls. Note the trends of increased neutrophil and decreased T cell proportions. b) Heatmap of changes in cell proportions of all datasets: non-COVID-19 *and* COVID-19 c) Concordant and discordant changes in cellular proportions comparing COVID-19 to non-COVID-19 viral infections. Cell types that increased in COVID-19 (hence decreased in non-COVID-19) were CD56^bright^ NK cells, M2 macrophages, and total NK cells. Those that decreased in non-COVID-19 but increased in COVID-19 were CD56^dim^ NK cells, memory B cells, and eosinophils.

We observed an overall correlation of 0.493 (p=0.017) for change in cellular proportions in patients with COVID-19 compared to non-COVID-19 viral infections (**Figure 6c, Supplementary Table 3**), where all but 6 cell types changed in the same direction, though not all changes were statistically significant. We again observed increased neutrophil and decreased T cell counts in COVID-19 which is in line with a recent study that compared COVID-19 to the 2009 H1N1^20^.Cell types that increased in COVID-19 relative to non-COVID-19 were CD56^bright^ NK cells, M2 macrophages, and total NK cells. Those that decreased in non-COVID-19 relative to COVID-19 were CD56^dim^ NK cells, memory B cells, and eosinophils. Although change in memory B cells was not statistically significant, the direction of change is expected as patients with non-COVID-19 infection are highly likely to have memory to those viruses, whereas SARS-CoV-2 is a novel coronavirus with no pre-existing memory in the population. Similar findings are reported when the absolute cell counts were measured by flow cytometry in smaller patient populations^20^.

## Discussion

Understanding the pathophysiology of COVID-19 is critical to finding new treatments. Here we take a host response transcriptomics approach using peripheral blood transcriptomics of the immune response to COVID-19 (n=62) compared to 652 non-COVID-19 viral infections spanning 6 viruses. While the vast majority of the host immune response appears to be similar between COVID-19 and other viruses, our study highlights some key differences.

The scatter plot of the correlation of the differential expression of non-COVID-19 viral infections versus COVID-19 infections illustrates this large proportion of concordance and seemingly small amount of discordance (Figure 4). We found only two genes, *ACO1* and *ATL3*, that were expressed in opposite directions. *ACO1* was over-expressed in COVID-19 versus HC and under-expressed in non-COVID-19 viral infections versus HC, whereas *ATL3* entirely oppositely regulated (Figure 4). Prior reports suggest that both genes may be involved in viral replication and immune evasion. *ACO1* is an iron-sulfur protein that regulates ferritin and transferrin. When cellular iron levels are low, the protein binds to iron-responsive elements (IREs), which represses translation of ferritin (a protein that stores iron), and simultaneously stabilizes the normally rapidly degraded transferrin receptor mRNA allowing for translation of the receptor and more cellular uptake of iron, which is required for proliferation^45^. High levels of ferritin are also indicative of macrophage activation syndrome (MAS) and have been observed in COVID-19 patients ^22,46–48^. *ATL3* is a member of the integral membrane GTPases. Proper formation of ER tubules is affected by mutations in this gene. Viruses are known to target host organelles to enter a host cell and avoid destruction^49^. Lack of ATL results in delayed cargo exit and coat assembly for budding from the ER which is necessary for export of cytokines and chemokines in response to infection; *ATL3* has been linked directly to viral replication in Zika^44^, although Zika was not studied here.

The power of using COCONUT to combine heterogeneous datasets allowed for a pooled, head-to-head comparison of COVID-19 with non-COVID-19 viral infections. Interestingly, the differentially expressed genes in this analysis were not enriched for any GO terms. However, there is bias in the annotation of gene ontologies, so absence of evidence does not denote evidence of absence of coordinated differential response ^50,51^. Indeed, **Figure 5** illustrates the comparison of COVID-19 to non-COVID-19 GO terms. We found many downregulated pathways are discordant when comparing to healthy controls. Within these, a cluster of pathways that are high in COVID-19 and low in non-COVID-19 viral infections involve ribosome related processes. In SARS-CoV-1 infections it was determined that viral nsp1 disrupts ribosomal function^52^. The inverse cluster of pathways that are high in non-COVID-19 viral infections and low in COVID-19 positively regulate cell-cell adhesion, cell activation, leukocyte activation, immune response-activating cell surface receptor signaling, perhaps suggesting a more dysregulated immune response. While the host response to SARS-CoV-2 in essence is highly similar to other viral infections, it does clearly have some molecular differences. Of particular interest was the observation that while both diseases had increased type-1 interferon signaling pathways, the magnitude of this pathway response was lower in the COVID-19 **(Figure 5)**.

Interestingly, the consistency in the change in the immune cell proportions are mostly consistent across COVID-19 and non-COVID-19 datasets. Our results are in line with several recent studies that found high neutrophil-lymphocyte-ratio (NLR) in COVID-19 patients^38–41^. Expansion of CD56^bright^ NK cells is common in many viral infections, as part of recognizing and killing virally infected cells while orchestrating adaptive immune responses^53^. Comparing patients with COVID-19 to HCs shows an increase in NK cells (**Figure 6a**), largely driven by the CD56^bright^ population. When compared to non-COVID-19 viral infections the increase in NK cell (via CD56^bright^ NK cell) proportion remains high in the COVID-19 infections. This phenomenon was also directly observed using mass spectrometry to measure cell abundance over time in COVID-19 patients, and when considering factors most explanatory in those that recovered the cells that were the most dynamic included CD56^dim^ NK cells^54^.

When comparing COVID-19 to non-COVID-19 viral infections, we see M1 macrophage proportions are similar to that of other viral diseases, but the elevated M2 response is discordant. M1 macrophages are pro-inflammatory and kill invaders, whereas M2 macrophages are considered anti-inflammatory and reparative. A large body of work in bacterial sepsis found that individuals with high M1 profiles had increased mortality whereas those with a more evenly balanced M1/M2 were more likely to survive^55^. However, in general, monocytotropic viruses including SARS-CoV-1 have evolved mechanisms to interfere with effective macrophage polarization, favoring the M2 population for immune evasion. For example, virus-induced macrophage depletion is executed by viruses that carry pro-antiapoptotic proteins, thus initially reducing the number of M1s to skew population to M2 and avoid attack, then further suppress the production and action of type I IFNs, stunting the progression of M1 macrophage polarization^56^. This shift we see in the proportion of M2 macrophages in COVID-19 versus non-COVID-19 viral infections indicate that this novel pathogen may be executing these immune evasion techniques with a high degree of success.

Our study has some limitations due to the design of using public data for non-COVID-19 comparison. First, due to the limited nature of clinical studies in a pandemic, we had just 62 patients with COVID-19 compared to >650 with other viral infections, creating class imbalance in their comparison. Second, we did not investigate effects of severity on host response as this was mostly unavailable. It is possible that differences in severity between this COVID-19 cohort and the other viral cohorts was a confounder in our analysis. Third, we analysed differential expression at single pre-set significance and effect size thresholds. Choosing different thresholds (e.g., thresholds based on 80% statistical power in each analysis) would have identified different sets of differentially expressed genes. We provide ES and FDR values for all genes (**Supplementary Table 2**) to enable re-analysis of these genes based on thresholds that others may deem more appropriate. **Supplementary Figure 4** is also provided to show the GO term enrichment results by varying cut-offs.

## Conclusions

We here provide bulk RNAseq profiling of peripheral blood in COVID-19 in comparison to healthy controls which we derived a signature of 2002 genes for investigation of the biology and potentially pathophysiology of this disease. We compiled an extensive database of non-COVID-19 viral infections across many platforms, ages, diseases and locations globally to compare to healthy controls using metaintegration to derive a set of 635 genes representing the host response to known viral pathogens. We then used COCONUT to conormalize all of the data and directly compare COVID-19 to non-COVID-19 viral infections resulting in a signature of 416 gene. We used all of these analyses to identify both the similarities and differences in the underlying host response. While we identified that a large proportion of the host response is similar to that of other infections, we also identified key differences in individual genes, pathways, and cellularity that are suggestive of the clinical differences observed in COVID-19. Of particular interest are the potential roles of ACO1 and ATL3 in describing the differential host response in COVID-19 complemented by the 416 genes that may identify novel biology and further the understanding of ACO1 and ATL3, but our findings will need to be replicated in further clinical studies.

## Data Availability

The publicly available cohorts are available under their respective study IDs within the manuscript. The COVID-19 cohort has been deposited in GEO for public reuse (GSE152641).

## Acknowledgements

We are grateful to Ashley Prasse Miller, Mario Esquivel, and Oliver Liesenfeld of Inflammatix Clinical Affairs team for clinical sample availability, and Luciano Brocchieri and Silvia Tomaletti of TB-SEQ for helpful discussion.

## Author contributions

TES, YDH, PK, and EJGB designed the study; GND, JK, GV, EG, EK, GD, NA and EJGB conducted clinical studies; SAT, YDH, YH, SS, RP, JT, PK performed bioinformatics analysis; DR, MR, and SC processed samples; SAT, YDH, PK, and TES wrote the manuscript; all authors critically revised and approved the manuscript.

## Data availability

The public cohorts are available under their respective study IDs. The COVID-19 cohort is deposited in GEO for public reuse (GSE152641).

